# Physical activity and life expectancy in Queensland, Australia: a lifetable analysis

**DOI:** 10.64898/2026.07.17.26358309

**Authors:** Mary N. Wanjau, Stephanie L. Duncombe, Jade Kubler, Gabriel Dillon, Gregore I. Mielke, J. Lennert Veerman

**Author notes:** (Corresponding author) Griffith University, Gold Coast, Queensland, Australia.

## Abstract

**Objectives:** To estimate the life expectancy gains that could be realised from increases in Queenslanders’ physical activity (PA) levels.

**Design:** Lifetable analysis

**Setting, Participants:** We modelled the 2025 Queensland population aged ≥40 years.

**Modelled scenarios:** We applied two approaches. In the first, we estimated life expectancy differences between device-measured PA quartiles, with quartile 1 representing the least active and quartile 4 the most active. In the second, we compared observed device-measured PA levels in Queensland with scenarios in which all individuals moved to either ≥12,000 steps/day or ≤2,000 steps/day. We converted the steps per day by age group and PA quartile into equivalent daily minutes of moderate-intensity walking at 4.8 km/h. Additional scenarios were explored in sensitivity analyses.

**Main outcomes:** Changes in life expectancy, and total life-years gained over the lifetime of the modelled population. Benefits were also translated into minutes of life gained per additional hour walked.

**Results:** If all Queenslanders aged ≥40 years were as active as the most active quartile, life expectancy at birth could be 88.3 years, an increase of 4.8 years above the life expectancy at observed activity levels. The life expectancy differences between individuals in the least active quartile and the most active quartile was 9.7 years. Achieving the activity level of the most active quartile would require individuals in the lowest activity quartile to undertake an additional 85.9 minutes/day of moderate-intensity walking, with each extra hour of PA associated with an average gain of approximately 3 hours (177 minutes) of life. In step-based modelling, life expectancy in the most active scenario (all achieving ≥12,000 steps/day) was higher by ~7.1 years compared with the least active scenario (all at ≤2,000 steps/day).

**Conclusions:** Increasing PA could yield meaningful gains in life expectancy for Queenslanders, with the largest gains seen in least active individuals. Our findings strengthen the case for prioritising investment in PA-promoting programs and environments.

**Plain language summary:** *The known:* Regular physical activity is linked to better health and longer life. Device-based physical activity shows stronger links with health outcomes than self-reported activity.

*The new:* Using device-measured activity data and life table modelling, we found that Queensland adults aged ≥40 years with the highest activity levels lived almost 10 years longer than the least active. Each additional hour of moderate-intensity walking was associated with an estimated gain of 3 hours of life.

*The implications:* Increasing physical activity, particularly among the least active, could substantially improve population health and longevity. Policies and environments supporting safe, accessible, activity should be prioritised.

## Introduction

Regular physical activity (PA) is associated with health benefits across the lifespan, with low levels of PA linked to increased risk of premature mortality and a range of chronic diseases [1–4]. Advances in measurement have improved the accuracy of PA assessment and showed stronger dose–response relationships than previously observed with self-report questionnaires [1, 3]. Evidence from device-based studies indicates that all intensities of movement, including light-intensity and accumulated number of steps, might contribute to overall health [5]. Increases in activity yield the greatest health gains among inactive individuals, with gains continuing at higher activity levels [2, 3, 5].

Physical inactivity is estimated to cost the Australian health system AUD 2.4 billion annually, in addition to up to AUD 15.6 billion in productivity losses [6–8]. Nationally, more than one third of Australian adults aged 18–64 years and over, and half of those aged 65 years and older, do not meet PA guidelines. Inactivity is more prevalent in socioeconomically disadvantaged and regional areas [9]. Queensland, Australia’s second largest state by area, has a widely dispersed population and reflects these inequities [8, 9].

Using device-measured PA, prospective cohort analyses from the UK Biobank and cohorts in the United States (US), Norway, and Sweden show that PA has an impact on longevity [10, 11]. In the US, we showed that average life expectancy could be 4 to 6 years higher if all adults were as active as the top 25% of the population [12]. The least active quarter could gain up to 11 years by increasing their activity to the level of the most active 25%, and an additional hour of walking was associated with several hours of extra lifetime [12]. Comparable estimates for Australia are not available. This study provides the first Queensland-specific estimates of life expectancy gains that could be realised from increased PA.

## Methods

### Overview

We created a life table of the Queensland population aged ≥40 years using population numbers from June 2025 and mortality rates for 2024 from the Australian Bureau of Statistics (ABS) [13, 14]. We re-analysed accelerometry data from the HABITAT (How Areas in Brisbane Influence healTh and AcTivity) longitudinal cohort study to estimate the levels of total PA (steps per day). We modelled the relationship between PA and mortality using two approaches: one based on PA quartiles with corresponding hazard ratios [15], and one based on step count categories with corresponding hazard ratios [2]. We applied potential impact fraction calculations to vary mortality as a function of population PA levels [16]. We then derived life tables to estimate life expectancy at different levels of total PA.

#### Total physical activity data for Queensland

We re-analysed data from HABITAT participants who took part in a 2014 sub-study (N = 612; Mean age 60.6 years [SD 6.9; range 48-73]) [17]. Participants wore a triaxial accelerometer (ActiGraph wGT3X-BT) on their non-dominant wrist for seven days. Data were analysed using the GGIR package in R an open-access, internationally recognised tool that enables the derivation of accurate and standardised estimates of PA exposure [18]. PA data were analysed across three age groups: 48–54 years, 55–64 years, and 65–73 years with levels for the 48–54-year age group applied to individuals aged 40–54 years, and levels from the 65–73-year age group were applied to those aged 65–100 years.

### Lifetable analysis

#### Approach based on physical activity quartiles

The first analytical approach is based on the Ekelund, Tarp [15] pooled analysis of eight large international cohort studies which provides hazard ratios by PA quartile with the least active as reference [15] (Table 1). We analysed average daily step counts (total PA) and categorised PA by quartiles, with quartile 1 representing the least active and quartile 4 the most active. For ease of interpretation, we converted the steps per day by age group and PA quartile into equivalent daily minutes of moderate-intensity walking at 4.8 km/h (3.0 mph) [19]. We used a cadence-based approach where moderate-intensity walking at approximately 4.8 km/h speed was assumed to correspond to ~100 steps per minute [20].

**Table 1:**
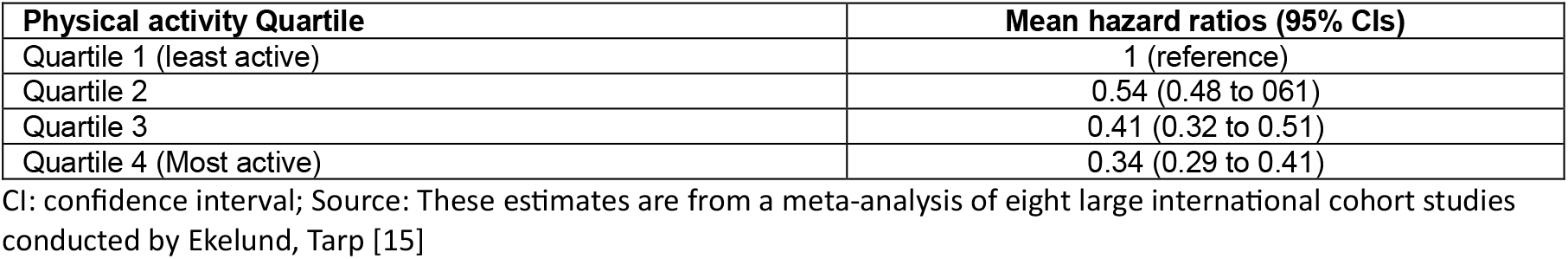
Pooled hazard ratios for the association between total physical activity by quartiles and all-cause mortality.

| Physical activity Quartile | Mean hazard ratios (95% CIs) |
| --- | --- |
| Quartile 1 (least active) | 1 (reference) |
| Quartile 2 | 0.54 (0.48 to 0.61) |
| Quartile 3 | 0.41 (0.32 to 0.51) |
| Quartile 4 (Most active) | 0.34 (0.29 to 0.41) |

CI: confidence interval; Source: These estimates are from a meta-analysis of eight large international cohort studies conducted by Ekelund, Tarp [15]

We used the ‘relative risk shift’ PIF calculation in which we changed the relative risk of the categories (PA quartiles) while keeping the proportion in each category constant (Equation 1) [16, 21]. Changes in all-cause mortality associated with different levels of PA modify age-specific probabilities of death, which in turn influence projected life-years lived and life expectancy estimates for the population. In our main results, we reported results for the life expectancy gains (in years) achieved by taking the difference in outcomes between 1) observed PA and each PA quartile and 2) moving up by one or more quartiles, covering all possible increases from adjacent to moving from lowest to the highest quartile.

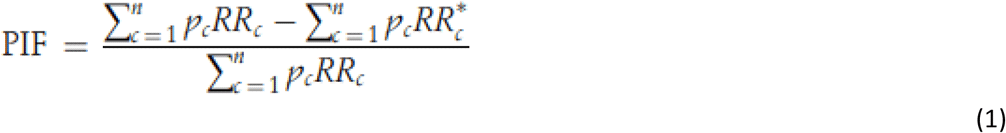

*Where Pc* is the proportion of the population in category c, *RRc* is the relative risk for that category, and *RRc\** is the relative risk of category c after the intervention.

For this approach, we also framed the life gains from the modelled scenarios as minutes of life gained (prolonged life) per additional hour of walking among adults aged ≥40 years. This was done by simulating the age-specific hours walked per year (365.25 days) by the population aged ≥40 years and deriving the additional hours required to move to higher PA quartiles (based on the required additional steps, converted into equivalent minutes of moderate-intensity walking at 4.8 km/h, as described earlier). Using these additional hours and the corresponding life years gained from movement across PA quartiles, we calculated the minutes gained (prolonged life) per additional hour of walking for each transition/scenario.

#### Approach based on physical activity in steps-per-day categories

The second approach is based on a meta-analysis by Ding, Nguyen [2] that provides pooled hazard ratios per 1,000-step increments (Table 2). We modelled PA as daily step counts across twelve categories ranging from ≤2,000 to 12,000+. For each category, we calculated the number and percentage of individuals achieving those levels, presented as frequency tables.

**Table 2:**
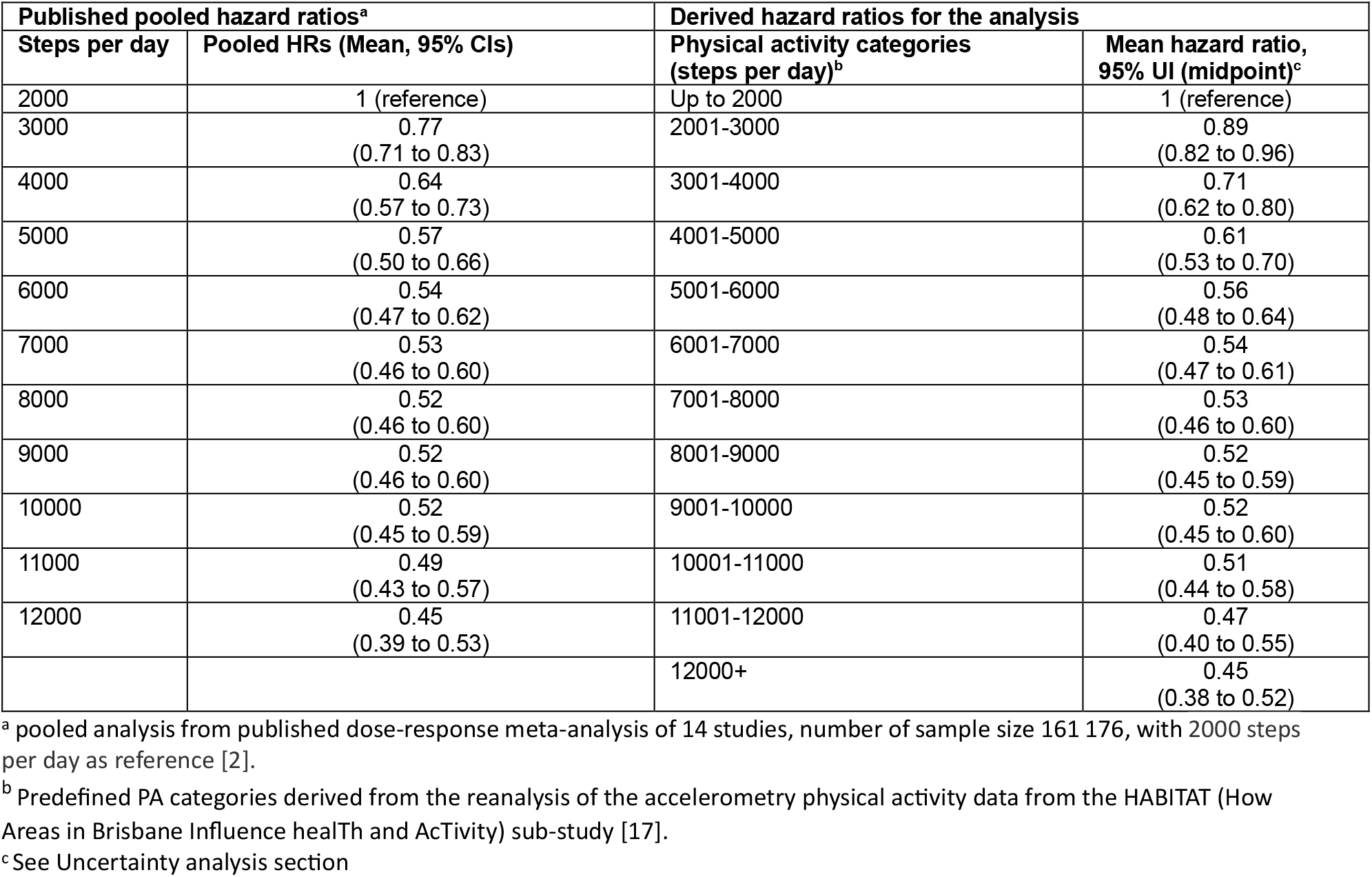
Hazard ratios used in the physical activity, steps per day modelling analysis.

| Published pooled hazard ratios<sup>a</sup> |  | Derived hazard ratios for the analysis |  |
| --- | --- | --- | --- |
| Steps per day | Pooled HRs (Mean, 95% CIs) | Physical activity categories (steps per day)<sup>b</sup> | Mean hazard ratio, 95% UI (midpoint)<sup>c</sup> |
| 2000 | 1 (reference) | Up to 2000 | 1 (reference) |
| 3000 | 0.77<br>(0.71 to 0.83) | 2001-3000 | 0.89<br>(0.82 to 0.96) |
| 4000 | 0.64<br>(0.57 to 0.73) | 3001-4000 | 0.71<br>(0.62 to 0.80) |
| 5000 | 0.57<br>(0.50 to 0.66) | 4001-5000 | 0.61<br>(0.53 to 0.70) |
| 6000 | 0.54<br>(0.47 to 0.62) | 5001-6000 | 0.56<br>(0.48 to 0.64) |
| 7000 | 0.53<br>(0.46 to 0.60) | 6001-7000 | 0.54<br>(0.47 to 0.61) |
| 8000 | 0.52<br>(0.46 to 0.60) | 7001-8000 | 0.53<br>(0.46 to 0.60) |
| 9000 | 0.52<br>(0.46 to 0.60) | 8001-9000 | 0.52<br>(0.45 to 0.59) |
| 10000 | 0.52<br>(0.45 to 0.59) | 9001-10000 | 0.52<br>(0.45 to 0.60) |
| 11000 | 0.49<br>(0.43 to 0.57) | 10001-11000 | 0.51<br>(0.44 to 0.58) |
| 12000 | 0.45<br>(0.39 to 0.53) | 11001-12000 | 0.47<br>(0.40 to 0.55) |
|  |  | 12000+ | 0.45<br>(0.38 to 0.52) |

<sup>a</sup> pooled analysis from published dose-response meta-analysis of 14 studies, number of sample size 161 176, with 2000 steps per day as reference [2].
<sup>b</sup> Predefined PA categories derived from the reanalysis of the accelerometry physical activity data from the HABITAT (How Areas in Brisbane Influence health and Activity) sub-study [17].
<sup>c</sup> See Uncertainty analysis section

We used the ‘proportions shift’ PIF calculation in which we changed the proportion of the Queensland population per defined PA category (Equation 2) [16]. Similar to the first simulation, changes in all-cause mortality associated with different levels of PA modify age-specific probabilities of death, which in turn influence projected life-years lived and life expectancy estimates for the population. To derive our results, we compared a Queensland population with the observed PA levels with 1) a scenario where all Queensland population moved to 12,000+ steps per day category (highest reported PA level) and 2) all Queensland population moved to the least active category (up to 2000 steps per day). We included additional scenarios in our sensitivity analysis: all moved to over 10,000 steps/day (those in over 10,000 retained their observed activity levels) and all moved to over 8,000 steps/day (all over 8,000 retained their observed activity levels). The model outputs included changes in life years over the lifetime of the 2025 Queensland population and life expectancy changes as the population moves from different levels of PA.

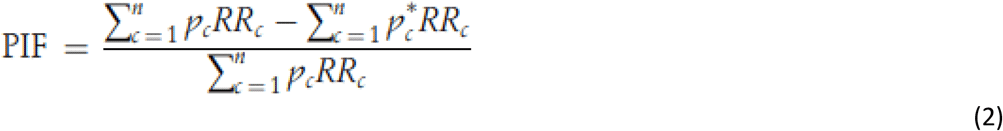

### Uncertainty analysis

The modelling analyses were implemented in MS Excel (Microsoft Office 365), 10,000 iterations, using the Ersatz add-in tool (EpiGear, Version 1.3) for uncertainty analysis [22]. The Ersatz tool uses Monte Carlo simulation to sample specified parameters for which probability distributions are defined within the model. We used the ErRelativeRisk function for the hazard ratios [23]. For Ding et al. hazard ratios [2], we derived the standard errors of the log risk ratios from the reported uncertainty intervals. We used the ‘ErRelativeRisk’ function in Ersatz software to incorporate parameter uncertainty [22, 23] and generated random deviates on the log scale, which were exponentiated to obtain midpoint hazard ratio model inputs. For PA by quartiles, we applied a normal distribution to average total PA levels to account for uncertainty in the model. Uncertainty in the PA category proportions was modelled using a Dirichlet distribution [22]. The uncertainty is propagated through the model to generate distributions of all model outputs, from which we report the mean and 95% uncertainty intervals (representing the 2.5^th^ and 97.5^th^ percentiles). We used the Guidelines for Accurate and Transparent Health Estimates Reporting (GATHER) in our reporting [24].

### Ethics

HABITAT received ethical clearance from the Queensland University of Technology Human Research Ethics Committee (ID numbers: 3967H & 1300000161).

## Results

### Physical activity levels from the HABITAT study

Overall, mean daily step counts ranged from 4,486 steps/day in the least active PA quartile (Q1) to 13,224 steps/day in the most active quartile (Q4) (Table 3). Across the PA quartiles, there was little difference in mean step counts (and equivalent daily minutes of walking) between age groups. Further, in the least and most active quartiles, overall activity was seen to decrease with age. Similarly, step-count distributions showed small differences by age (Table 4 and Table S1). Older adults (65+ years) were slightly more prevalent in lower-range step categories and had lower percentages achieving ≥12,000 steps/day when compared to younger adults.

**Table 3:**
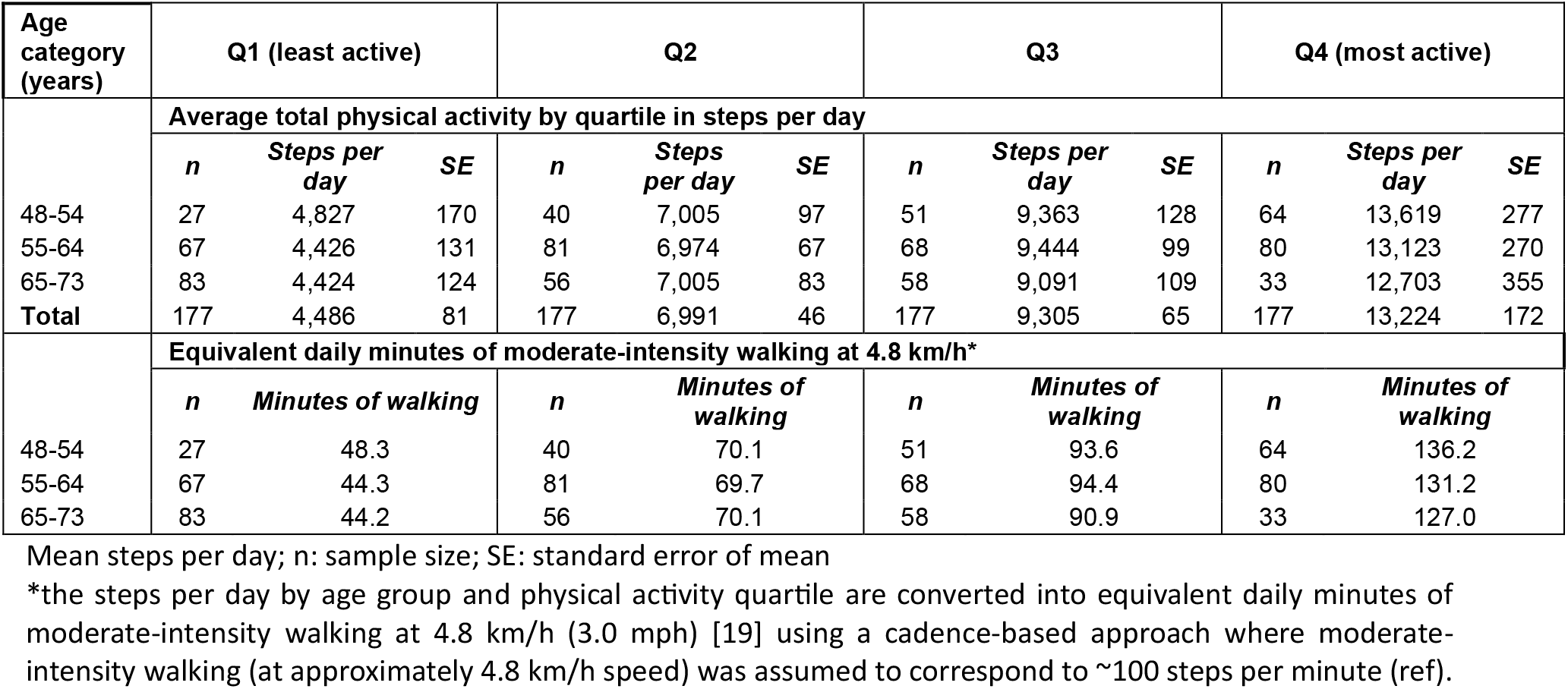
Average total physical activity from the HABITAT study by quartile in steps per day.

**Table 4:** Total physical activity in steps per day as percentage distributions.

| Steps per day | 48-54 years | 55-64 years | 65-73 years | Total sample |
| --- | --- | --- | --- | --- |
|  | **Total** | **Total** | **Total** | **Total** |
| Up to 2000 | 0% | 0% | 1% | 1% |
| 2001-3000 | 1% | 2% | 3% | 2% |
| 3001-4000 | 1% | 6% | 6% | 5% |
| 4001-5000 | 6% | 6% | 11% | 8% |
| 5001-6000 | 7% | 8% | 16% | 10% |
| 6001-7000 | 11% | 14% | 10% | 12% |
| 7001-8000 | 12% | 14% | 14% | 13% |
| 8001-9000 | 10% | 7% | 12% | 9% |
| 9001-10000 | 8% | 8% | 7% | 8% |
| 10001-11000 | 10% | 10% | 7% | 9% |
| 11001-12000 | 9% | 10% | 5% | 8% |
| 12000+ | 25% | 15% | 7% | 15% |
| Total | 100% | 100% | 100% | 100% |

### Changes in life expectancy: quartiles analysis

In our lifetable model, average life expectancy in Queensland in 2025 was estimated at 83.6 years (Figure 1). If all Queenslanders over the age of 40 were as active as the top 25% (Q4), life expectancy at birth would be 88.3 years (95% Uncertainty Interval [UI]: 86.9 to 89.7), which is an increase of 4.8 years (95% UI: 3.4 to 6.3) (Figure 1 and Table S2). If all Queenslanders in 2025 aged 40 years and above were as active as the least active 25% (Q1), life expectancy at birth would be 78.7 years (95% UI: 78.2 to 79.2), a projected loss of 4.9 years (95% UI: 4.4 to 5.4). If all Queenslanders in 2025 had PA at the levels corresponding to Q2 and Q3, a life expectancy at birth of 84.1 (95% UI: 83.2 to 85.0) and 86.4 (95% UI: 84.9 to 88.1) would be expected, respectively (Figure 1 and Table S2).

**Figure 1:**
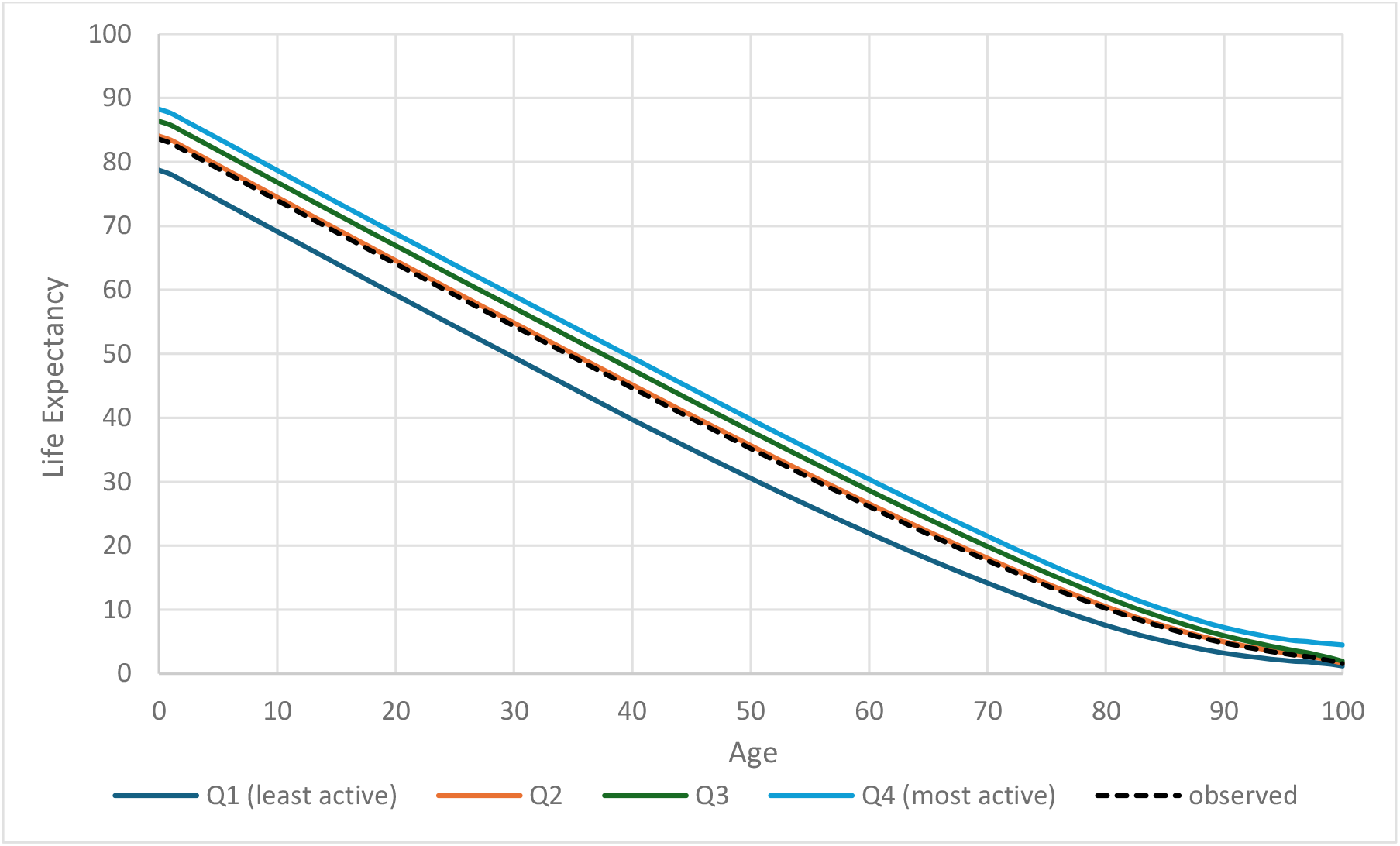
Queensland life expectancy by age, comparing observed to results by adult physical activity quartile

Life expectancy gains were realised when lower active individuals in the model moved to higher PA levels (Table 5 and Table S3). The life expectancy differences between individuals in the least active quartile and the most active quartile was 9.7 years (95% UI: 8.1 to 11.4). The non-linear risk curve, which is steep at lower levels of PA (Table 1), translates into a pattern of diminishing returns, with the greatest differences observed between the least active quartile and Q2 – a difference of 5.4 years (95% UI: 4.4 to 6.5) (Table 5). Achieving the health benefits of the most active quartile would require individuals in the least active quartile to undertake the equivalent of an extra 85.9 minutes/day (95% UI: 82.1 to 89.7) of walking (4.8 km/h) per person. For this transition from least to most active quartile, on average, each additional hour of PA would prolong life by 176.8 life-minutes (95% UI: 153.9 to 199.6) (Table 5).

**Table 5:** Benefits achieved by lower active individuals when they move to higher physical activity levels in the model.

| Change in physical activity (quartile) | Average extra 4.8 km/h walking equivalence (minutes per day) | Minutes gained (prolonged life) per additional hour walked above age 40 | Life-expectancy gain (years) |
| --- | --- | --- | --- |
| 1 → 2 | 24.1 (22.4 to 25.8) | 542.5 (445.9 to 641.0) | 5.4 (4.4 to 6.5) |
| 1 → 3 | 47.0 (45.1 to 48.9) | 306.3 (244.3 to 364.7) | 7.9 (5.9 to 9.8) |
| 1 → 4 | 85.9 (82.1 to 89.7) | 176.8 (153.9 to 199.6) | 9.7 (8.1 to 11.4) |
| 2 → 3 | 22.9 (21.1 to 24.6) | 111.9 (12.1 to 208.0) | 2.4 (0.2 to 4.6) |
| 2 → 4 | 61.8 (58.1 to 65.5) | 53.6 (26.8 to 80.4) | 4.3 (2.4 to 6.2) |
| 3 → 4 | 38.9 (35.1 to 42.7) | 13.3 (-44.4 to 75.9) | 1.9 (-0.6 to 4.5) |

Results for the additional hours walked per year from movement to higher physical activity quartiles and corresponding life years gained from movement across physical activity quartiles are presented in Table S3.

### Changes in life expectancy: steps per day analysis

If the entire modelled population achieved ≥12,000 steps per day (most active category), this could increase life expectancy by 1.7 years (95% UI: 1.6 to 1.9) (Table 6 and Table S4). If the entire modelled population was in the least active category (≤2,000 steps per day), this could reduce life expectancy by 5.4 years for the total population (95% UI: −6.6 to −4.1).

**Table 6:**
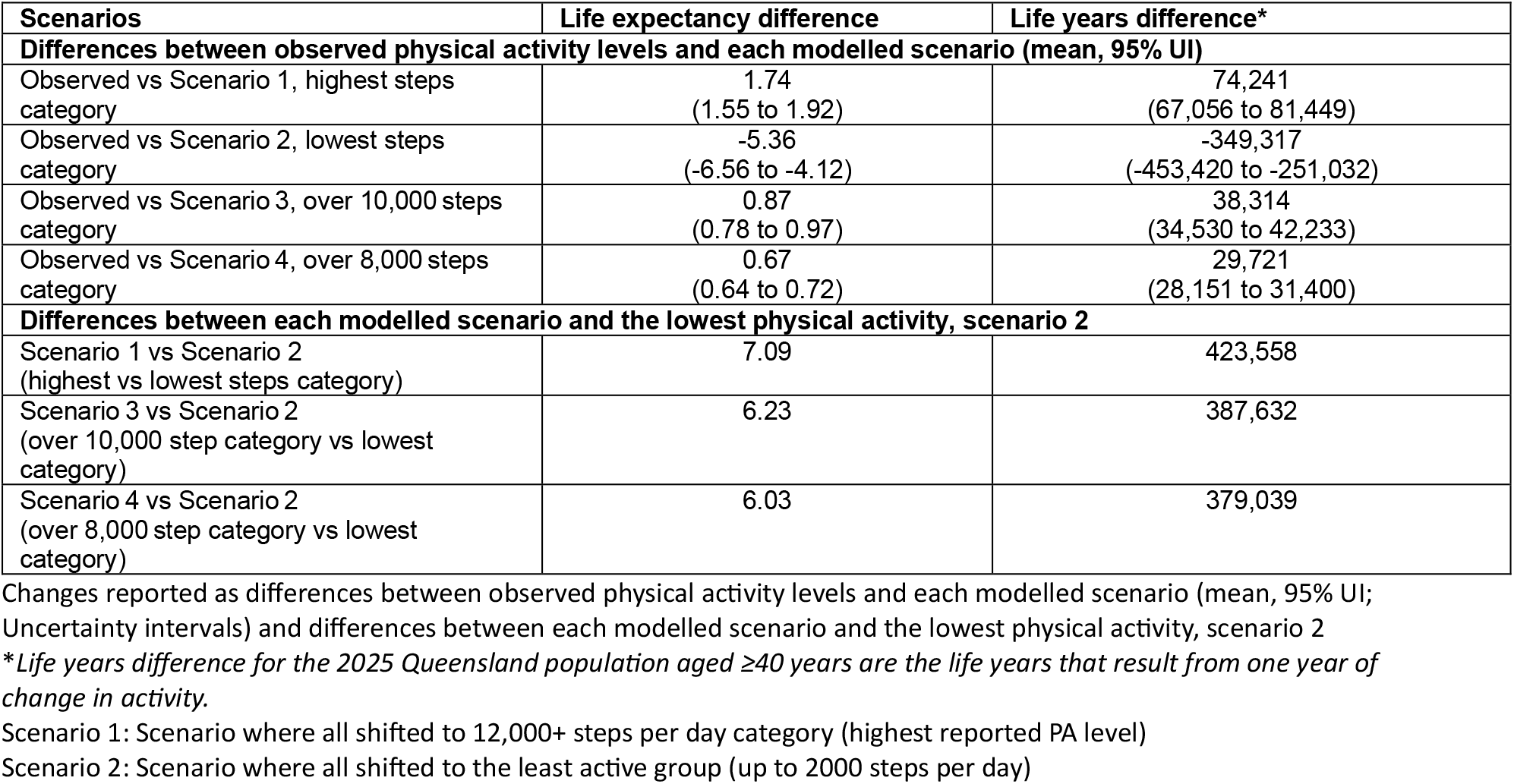
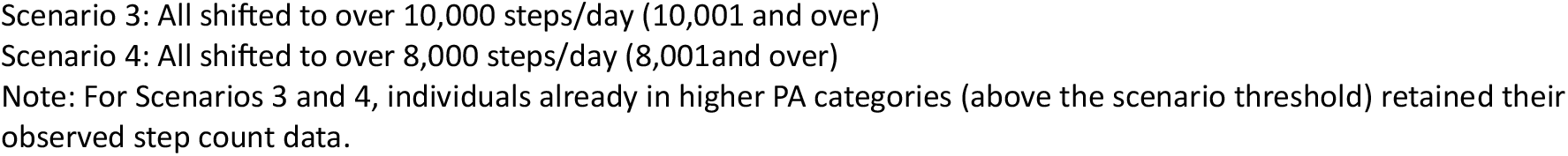
Changes in life expectancy and life years of the 2025 Queensland population aged ≥40 years.

| Scenarios | Life expectancy difference | Life years difference* |
| --- | --- | --- |
| **Differences between observed physical activity levels and each modelled scenario (mean, 95% UI)** |  |  |
| Observed vs Scenario 1, highest steps category | 1.74<br>(1.55 to 1.92) | 74,241<br>(67,056 to 81,449) |
| Observed vs Scenario 2, lowest steps category | -5.36<br>(-6.56 to -4.12) | -349,317<br>(-453,420 to -251,032) |
| Observed vs Scenario 3, over 10,000 steps category | 0.87<br>(0.78 to 0.97) | 38,314<br>(34,530 to 42,233) |
| Observed vs Scenario 4, over 8,000 steps category | 0.67<br>(0.64 to 0.72) | 29,721<br>(28,151 to 31,400) |
| **Differences between each modelled scenario and the lowest physical activity, scenario 2** |  |  |
| Scenario 1 vs Scenario 2 (highest vs lowest steps category) | 7.09 | 423,558 |
| Scenario 3 vs Scenario 2 (over 10,000 step category vs lowest category) | 6.23 | 387,632 |
| Scenario 4 vs Scenario 2 (over 8,000 step category vs lowest category) | 6.03 | 379,039 |

Changes reported as differences between observed physical activity levels and each modelled scenario (mean, 95% UI; Uncertainty intervals) and differences between each modelled scenario and the lowest physical activity, scenario 2
\*Life years difference for the 2025 Queensland population aged $\geq 40$ years are the life years that result from one year of change in activity.
Scenario 1: Scenario where all shifted to 12,000+ steps per day category (highest reported PA level)
Scenario 2: Scenario where all shifted to the least active group (up to 2000 steps per day)
Scenario 3: All shifted to over 10,000 steps/day (10,001 and over)
Scenario 4: All shifted to over 8,000 steps/day (8,001 and over)
Note: For Scenarios 3 and 4, individuals already in higher PA categories (above the scenario threshold) retained their observed step count data.

We found that life expectancy in the most active scenario (all individuals achieving ≥12,000 steps/day) was higher by ~7.1 years compared with the least active scenario (all individuals at ≤2,000 steps/day). The results for the changes in life expectancy and life years for all modelled scenarios are provided in Table 6.

## Discussion

If all Queenslanders over the age of 40 were as active as the most active quartile, life expectancy at birth could be 88.3 years, an increase of 4.8 years. The greatest life expectancy differences were seen between individuals in the least active quartile and the most active quartile with a 9.7-year difference. Achieving the health benefits of the most active quartile would require individuals in the lowest activity quartile to undertake the equivalent of an extra 85.9 minutes/day of walking (at 4.8 km/h). For this group, on average, each additional hour of walking would prolong life by ~3 hours (177 minutes).

In the second modelling approach, life expectancy in the most active scenario (all achieving ≥12,000 steps/day) was higher by ~7.1 years compared with the least active scenario. If all Queenslanders over the age of 40 years achieved ≥12,000 steps per day, life expectancy could increase by 1.7 years. This is a much smaller effect compared to the relative (quartile) approach because it applies a fixed absolute threshold, which limits improvements for individuals already near that level, while the quartile-based approach redistributes PA across the full population distribution, allowing broader cumulative shifts. In addition, the hazard ratios by Ding, Nguyen [2] show less steep risk gradients than those by Ekelund, Tarp [15] used in our first modelling approach. In this study, the data from the HABITAT study show a relatively active sample. Our model estimates suggest that much of the life expectancy gains from PA have already been realised by the HABITAT participants, leaving less to gain by moving more. Life expectancy in the highest activity scenario (all individuals achieving ≥12,000 steps/day) was higher by ~7.1 years compared with the lowest PA scenario (all individuals at ≤2,000 steps/day).

Strengths of this study include generating evidence specific to Queensland State using the most representative objectively measured PA data that were available for the state, based on average daily step counts - an intuitive and easily tracked metric. We expanded our previous work [12] to include two approaches where PA is modelled using both quartiles and step-count categories improving interpretation across multiple audiences and strengthening evidence across plausible activity scenarios.

Several limitations deserve mention. The use of 2014 HABITAT data collected seven years after cohort recruitment is a limitation; results may have been affected by differential loss to follow-up. Generalisability is also limited due to the cohort’s concentration in Brisbane, a metropolitan area, where adults are likely more active than in regional areas [8]. Because these estimates rely on a single-point assessment of PA, they may underestimate the true associations and potential gains; studies using long-term assessments have consistently reported stronger associations with mortality risk [25]. Our current findings may thus underestimate potential gains in less active and regional populations. Another limitation is that to enable modelling across ages 40-100 years, PA levels from adjacent age groups were applied, which may have underestimated activity in younger adults (40-47 years) and overestimated activity in older adults (≥74 years) as PA generally declines with age [9].

Changes in Brisbane’s built environment since 2014, as well as broader social, demographic, and policy factors, may also impact current PA levels [26]. Our study estimates average longevity gains across PA quartiles. At the individual level, the capacity to increase PA varies. Finally, mortality risks were modelled using robust hazard ratios from large device-based cohort meta-analyses with extensive confounder adjustment and sensitivity analyses [2, 15] but residual confounding cannot be excluded.

### Comparison with other studies

Compared with our previous US estimates [12], the outcomes for Queensland are smaller. For the US, life expectancy increased by 10.9 years, and by 6.3 years when comparing the difference between individuals in the least active quartile and those in the most active quartile and between individuals in the least active quartile and those in the second quartile, respectively. In Queensland, life expectancy increased by 9.7 years, and by 5.5 years when comparing the difference between individuals in the least active quartile and those in the most active quartile and between individuals in the least active quartile and second quartile, respectively. This is likely due to the higher mortality rates in the US compared to Queensland. However, the average gain per additional hour of activity was similar in both populations (approximately three extra hours of life per hour of activity).

In a prospective cohort analysis of UK Biobank participants with objectively measured PA, an additional daily 10-min brisk walk was associated with a life expectancy increase of 0.9 years in inactive women and 1.4 years in inactive men [11]. Our model suggests a 10-minute walk at 4.8 km/h increases life expectancy by 1.1 years for both sexes combined when comparing individuals in the least active quartile and those in the most active quartile and by 2.2 years when comparing individuals in the least active quartile and those in second quartile. Our estimates reflect a pattern of diminishing returns due to non-linear risk curve. The UK Biobank estimates reflect individual-level gains among inactive participants, whereas our estimates represent population-level average effects across PA quartiles. A meta-analysis of UK Biobank and cohorts in Norway, Sweden, and the US, found that small increases of 5 min/day of moderate-to-vigorous PA among the least active participants could prevent approximately 6% of all deaths [10]. Similar to our study, these authors found that modest achievable increases in PA, particularly among the least active, can result in substantial gains in longevity.

### Policy relevance and implications and future research

By demonstrating that increased PA is likely to improve life expectancy, particularly among the least active, these findings support PA as a priority for health policy and practice. Population-level policies addressing structural and social barriers can enable modest achievable increases in PA equitably, with priority given to inactive populations particularly given that inactivity is more prevalent in socioeconomically disadvantaged and regional areas (12). In Queensland, these could include PA promoting infrastructure, policies, and programmatic and social prescribing responses, walkable neighbourhoods, accessible public transport and community programs [27], and leveraging the 2032 Olympic and Paralympic Games to deliver a lasting and inclusive health legacy [28].

Modelling PA using daily step count categories aligns with how people monitor activity and reflects evolving policy practice, including the recent incorporation of step-based guidance in companion statements supporting the latest Australian 24-Hour Movement Guidelines [29]. Recognising that health benefits accrue from all forms of PA, framing PA in terms of steps per day provides a practical way of contextualising moderate-intensity PA recommendations, strengthening links between national guidelines and supporting translation of guideline targets into everyday activity patterns.

Future research could extend analyses to estimate health and economic gains by socioeconomic status, rural–urban location, and other disadvantaged groups, and to younger adults aged 18– 40 years. This would require the collection of representative accelerometry data in these groups. There is also a strong case for population-specific assessments among people living with chronic conditions, particularly in the context of an ageing population where multimorbidity is common. Research could also assess how changes in urban and rural infrastructure influence PA levels and evaluate these investments to determine their health, economic and equity impacts and cost-effectiveness [30].

## Conclusion

Increasing PA in Queensland could yield meaningful gains in life expectancy. The greatest benefits were seen in the least active individuals. By using device-measured data and translating results into life expectancy gains per additional hour of walking, the findings provide clear, intuitive policy-relevant evidence that can strengthen the case for prioritising equitable, population-wide investment in physical-activity-promoting environments and policies across Queensland.

## Funding

Health and Wellbeing Queensland

## Conflicts of interest

JK and GD are employed by Health and Wellbeing Queensland, who provided funding for this study. All other authors declare no conflict of interest

## Acknowledgements

We would like to acknowledge and thank the HABITAT (How Areas in Brisbane Influence healTh and AcTivity) study participants. We would like to acknowledge our HABITAT Co-CIs and project staff for assistance with survey development, project management, data collection, coding and cleaning, and data management.

## Author contribution statement

**Mary N. Wanjau:** conceptualization, methodology, data curation, validation, writing – original draft, writing – review & editing, project administration, funding acquisition

**stephanie L. Duncombe:** data curation, writing – review & editing

**Jade Kubler:** writing – review & editing

**Gabriel Dillon:** writing – review & editing

**Gregore I. Mielke:** data curation, writing – review & editing

**J. Lennert Veerman:** conceptualization, methodology, validation, writing – review & editing, funding acquisition, supervision

## Data sharing statement

The study data are reported in the manuscript and supplementary file

## Notes

### Competing Interest Statement

The authors have declared no competing interest.

### Author Declarations

HABITAT study received ethical clearance from the Queensland University of Technology Human Research Ethics Committee (ID numbers: 3967H & 1300000161).

