## Supplementary File for "Physical activity and life expectancy in Queensland, Australia: a lifetable analysis"

**Authors**

Mary N. Wanjau

Griffith University, Gold Coast, Queensland, Australia

Stephanie L. Duncombe

The University of Queensland, Brisbane, Queensland, Australia

Jade Kubler

Health and Wellbeing Queensland, Brisbane, Queensland, Australia

Gabriel Dillon

Health and Wellbeing Queensland, Brisbane, Queensland, Australia

Gregore I. Mielke

The University of Queensland, Brisbane, Queensland, Australia

J. Lennert Veerman (Corresponding author)

Griffith University, Gold Coast, Queensland, Australia

### Sex specific physical activity levels from the HABITAT study

Table S1: Total physical activity in steps per day as percentage distributions

| Steps per day | 48-54 years | | | 55-64 years | | | 65+ years | | | Total sample | | |
|---|---|---|---|---|---|---|---|---|---|---|---|---|
| | Male | Female | Total | Male | Female | Total | Male | Female | Total | Male | Female | Total |
| Up to 2000 | 0% | 0% | 0% | 0% | 1% | 0% | 1% | 1% | 1% | 0% | 1% | 1% |
| 2001-3000 | 3% | 0% | 1% | 4% | 1% | 2% | 3% | 4% | 3% | 3% | 2% | 2% |
| 3001-4000 | 1% | 1% | 1% | 7% | 5% | 6% | 8% | 4% | 6% | 6% | 4% | 5% |
| 4001-5000 | 6% | 6% | 6% | 5% | 7% | 6% | 15% | 9% | 11% | 9% | 7% | 8% |
| 5001-6000 | 9% | 6% | 7% | 9% | 8% | 8% | 11% | 19% | 16% | 10% | 11% | 10% |
| 6001-7000 | 9% | 13% | 11% | 15% | 13% | 14% | 12% | 8% | 10% | 12% | 11% | 12% |
| 7001-8000 | 13% | 11% | 12% | 13% | 14% | 14% | 13% | 15% | 14% | 13% | 14% | 13% |
| 8001-9000 | 11% | 9% | 10% | 7% | 8% | 7% | 12% | 12% | 12% | 10% | 9% | 9% |
| 9001-10000 | 10% | 7% | 8% | 5% | 9% | 8% | 12% | 4% | 7% | 9% | 7% | 8% |
| 10001-11000 | 10% | 10% | 10% | 10% | 10% | 10% | 2% | 10% | 7% | 7% | 10% | 9% |
| 11001-12000 | 9% | 9% | 9% | 14% | 8% | 10% | 2% | 7% | 5% | 9% | 8% | 8% |
| 12000+ | 19% | 30% | 25% | 11% | 17% | 15% | 8% | 7% | 7% | 12% | 17% | 15% |
| Total | 100% | 100% | 100% | 100% | 100% | 100% | 100% | 100% | 100% | 100% | 100% | 100% |

### Additional results from the approach based on physical activity quartiles

*Table S2: Changes in life expectancy*

| Variable | Mean (years) | 95% Uncertainty Interval boundaries | |
|---|---|---|---|
| **Average life expectancy at birth (years)** | | | |
| Observed in Queensland | 83.6 | 83.6 | 83.6 |
| If all were as active as the least active 25% (Q1) | 78.7 | 78.2 | 79.2 |
| If all were as active as the Q2 PA level | 84.1 | 83.2 | 85.0 |
| If all were as active as the Q3 PA level | 86.4 | 84.9 | 88.1 |
| If all were as active as the top 25% (Q4) | 88.3 | 86.9 | 89.7 |
| **Life-expectancy at age 40 years** | | | |
| Observed in Queensland | 44.7 | 44.7 | 44.7 |
| If all in the least active 25% (Q1) | 39.7 | 39.2 | 40.2 |
| If all were as active as the Q2 PA level | 45.2 | 44.3 | 46.1 |
| If all were as active as the Q3 PA level | 47.6 | 46.0 | 49.2 |
| If all were as active as the top 25% (Q4) | 49.4 | 48.0 | 50.9 |
| **Changes in life expectancy** | | | |
| Difference between observed and the least active 25% (Q1) | - 4.9 | - 5.4 | - 4.4 |
| Difference between observed and Q2 | 0.5 | - 0.4 | 1.4 |
| Difference between observed and Q3 | 2.9 | 1.3 | 4.6 |
| Difference between observed and most active 25% (Q4) | 4.8 | 3.4 | 6.3 |
| Difference between the least active 25% (Q1) and Q2 | 5.4 | 4.4 | 6.5 |
| Difference between Q1 and Q3 | 7.9 | 5.9 | 9.8 |
| Difference between Q1 and most active 25% (Q4) | 9.7 | 8.1 | 11.4 |
| Difference between Q2 and Q3 | 2.4 | 0.2 | 4.6 |
| Difference between Q2 and Q4 | 4.3 | 2.4 | 6.2 |
| Difference between Q3 and Q4 | 1.9 | - 0.6 | 4.5 |

Results reported for both sexes combined. Q1 to Q4: average daily step counts (total physical activity) by quartile, with quartile 1 representing the least active and quartile 4 the most active
Observed physical activity is average daily step counts (total physical activity) for all of the Queensland population derived from the HABITAT study [1].

*Table S3: Additional hours walked per year and corresponding life years gained from movement across physical activity quartiles*

| Change in PA (quartile) | Hours walked per year (mean, 95% uncertainty interval) | Life years gained[#] (mean, 95% uncertainty interval) |
|---|---|---|
| **1 → 2** | 96,829,073 (89,490,360 to 104,157,772) | 99,722 (83,388 to 115,614) |
| **1 → 3** | 214,150,560 (205,649,156 to 222,812,534) | 124,649 (99,630 to 147,719) |
| **1 → 4** | 383,634,700 (367,596,344 to 399,959,616) | 128,922 (112,907 to 144,317) |
| **2 → 3** | 117,321,487 (110,097,247 to 124,524,083) | 24,927 (2,669 to 46,224) |
| **2 → 4** | 286,805,627 (271,457,457 to 302,676,392) | 29,200 (14,663 to 43,746) |
| **3 → 4** | 169,484,140 (153,268,206 to 185,832,857) | 4,273 (-14,223 to 24,295) |

[#]*Life years gained for the 2025 Queensland population aged ≥40 years: the life years that result from one year of change in activity.*

Life years difference, $\Delta Ly = \Sigma$ (delta qx * ex * lx), i.e., age-specific probability of dying at observed PA - probability of dying at the reference PA quartile * life expectancy at that age before intervention (assuming that after that first year, normal PA levels apply again) * QLD population size

### Additional results from the approach based on physical activity in steps-per-day categories

*Table S4: Life expectancy*

Sex-specific life expectancy for the observed physical activity levels and for each modelled scenario

| **Variable** | **Mean, 95% Uncertainty Intervals (years)** |
|---|---|
| **Average life expectancy at birth (years)** | |
| Observed in Queensland | 83.6 (83.6 to 83.6) |
| If the entire modelled population achieved ≥12,000 steps per day (most active category) | 85.3 (85.1 to 85.5) |
| If the entire modelled population was in the least active category (≤2,000 steps per day) | 78.3 (77.1 to 79.5) |
| If all moved to over 10,000 steps/day (10,001 and over) | 84.4 (84.3 to 84.5) |
| If all moved to over 8,000 steps/ day (8,001 and over) | 84.2 (84.2 to 84.3) |
| **Life-expectancy at age 40 years** | |
| Observed in Queensland | 44.7 (44.7 to 44.7) |
| If the entire modelled population achieved ≥12,000 steps per day (most active category) | 46.4 (46.2 to 46.6) |
| If the entire modelled population was in the least active category (≤2,000 steps per day) | 39.3 (38.1 to 40.5) |
| If all moved to over 10,000 steps/day (10,001 and over) | 45.5 (45.4 to 45.6) |
| If all moved to over 8,000 steps/ day (8,001 and over) | 45.3 (45.3 to 45.4) |

Results reported for both sexes combined
